# Quantifying additive and multiplicative effects of socially stigmatized identities on self-reported overall health

**DOI:** 10.1101/2025.02.06.25321809

**Authors:** Katrine LeStage, Robert Vogel, Julia Moore Vogel

**Affiliations:** Hoosic Valley Central School, High School, Schaghticoke, NY, United States; University of California, Department of Psychiatry, San Diego, La Jolla, CA, United States; Scripps Research, Translational Institute, La Jolla, CA, United States

**Author notes:** **Corresponding author:** Julia Moore Vogel, PhD, 3344 N Torrey Pines Ct, La Jolla, CA, 92037.

**Keywords:** health disparities, overall health, *All of Us* Research Program, intersectionality, public health

## Abstract

Individuals with one or more socially stigmatized identities experience extensive health disparities. However, most studies consider the effects of only individual stigmatized identities. Further, the effects of individual identities and their intersections on overall health have not been quantified. We used participant-reported survey data collected in the *All of Us* Research Program and released to the controlled tier in April 2023 to statistically estimate the first and second order effects of 47 stigmatized identities on self-reported overall health. After using false discovery rate to adjust for testing multiple hypotheses, 29 individual stigmas had statistically significant effects on self-reported overall health and 116 pairs of stigmas did. All significant individual effects were negative or neutral except for skin cancer. Those with the largest negative effect on self-rated overall health are difficulty walking or climbing stairs, unemployed or unable to work, difficulty with errands, and low educational attainment. Pairs of intersecting stigmas had a mix of negative and positive incremental effects, indicating that some stigmatized identities are negative modifiers, such as depression, and other combinations are less negative that the sum of their individual negative effects, such as having difficulty with multiple types of activities of daily living. Taken together, there are numerous pairs of stigmatized identities that significantly affect self-reported overall health and therefore should be considered in research and clinical care.

## Introduction

Individuals with socially stigmatized identities experience extensive health disparities ^1,2^. Individuals with multiple identities that are socially stigmatized report experiencing more discrimination and lower quality of life ^3,4^, reflecting the connection between intersectionality and health disparities ^5,6^. However, most studies of participants experiencing health disparities analyze each group in isolation; ignoring the multifaceted nature of each individual can lead to incorrect conclusions ^7^. Further, the effects of specific stigmatized identities, which we refer to below as stigmatizations, and their intersections have not been quantified.

Individuals who may be clinically evaluated to have equivalent overall health may self-report different overall health ratings ^8^, limiting the value of direct comparisons between groups. There are cultural differences and other population-specific biases with self-reported health data ^8–10^. Individuals may evaluate their health relative to their expectations, their lived experience, and those around them. In cases where an objective measure would report higher overall health than a self-reported one, the difference remains significant, as perception of one’s health can affect health outcomes ^11^.

To date concomitant quantitative estimates of additive and multiplicative effects has been lacking. A lack of the requisite sample size has precluded such analysis, given the need for large numbers (e.g. over a thousand) of simultaneous statistical estimates under a linear model. To this end, we utilize the large scale and diverse participant base of the *All of Us* Research Program ^12,13^ to analyze self-reported overall health from 387,411 participants within 47 individual and 1,124 pairs of stigmatized identities. We statistically estimate the additive and multiplicative effects of stigmatizations on self-reported overall health.

## Methods

We used *All of Us* Research Program participant survey responses^14^ to create cohorts for each single and pairwise stigma within the *All of Us* Researcher workbench controlled tier version 7^15^. Most stigmas had clear data types associated with them, for example race and ethnicity were identified using responses from the Basics survey and most health conditions were from the Personal and Family Health History Surveys. Overall Health ratings were from the Overall Health survey, in particular the question “In general, would you say your overall health is:” were converted to the integers Excellent, Very Good, Good, Fair, Poor. We chose to use survey data and not electronic health data because the surveys are likely taken within close proximity, whereas electronic health record data can span decades.

We used a linear model to estimate the effects of individual and pairwise stigmas on overall health self-ratings. Estimates of effect-sizes under the model were computed by the *lm* linear regression tool in the R statistical programming language. To ensure that the effect-size estimates are unique, we eliminated collinear stigmatizations so that the matrix of covariates was full rank. The two elements that were removed were Black and Hispanic or Latino (which is a subset of Black and Multiracial) and Asian and Hispanic or Latino (which is a subset of Asian and Multiracial). Cohorts were created using the *All of Us* Researcher Workbench’s cohort builder, data analysis was done within the Researcher Workbench in R, and figures were created in R Studio version 2024.09.0+375.

### Ethics Statement

The study was reviewed by the Scripps IRB and determined exempt human subjects research. The data were accessed between April 5, 2023 and November 15, 2024; through this access, the research team did not have access to data that could identify potential participants.

## Results

We compared *All of Us* Research Program participants’ self-reported overall health ^16^ within and between each individual stigmatizations and pairs of stigmatizations. We examined responses to questions regarding overall health, physical health, mental health, and quality of life (Figure S1), and found an average correlation of 0.61 between responses for a given individual. For simplicity and to preserve statistical power, we constrained further analysis to overall health scores.

Stigmatizations were defined based on a prior study defining 93 stigmas that affect health, including demographic categories, disability statuses, and various health conditions ^17^. All 47 that could be practicably included from *All of Us* Research Program survey data, with a sample size of at least 150 participants per stigma, were included in the analyses. First, we separately examined the average self-reported overall health within individual (Figure S2a) and pairs of stigmatizations (Figure S2b). Disability statuses (in particular, difficulty with activities of daily living) had the lowest average overall health self-ratings followed by low educational attainment. Asian, multiracial and Gay and/or Lesbian individuals as well as those with breast, bladder, or prostate cancer were amongst those with the highest average overall health self-ratings, all of which were higher than the average for the cohort as a whole (Figure S2a). Individuals without stigmatized identities had a higher self-reported overall health than any stigmatized identity. Due to relatively large sample sizes (Figure S3), most individual stigmatizations have mutually exclusive standard error of the mean.

Next, we created a model to simultaneously estimate only the effect of individual stigmatizations. Then, we created a model to simultaneously estimate the effect size of all single and pairwise stigmatizations on self-reported overall health. There are notable differences between the two models; for example difficulties with activities of daily living, low educational attainment, Intersex, Black, and Hispanic/Latino have stronger negative effects in the model that considers multiplicity (Figure 1).

**Figure 1.**
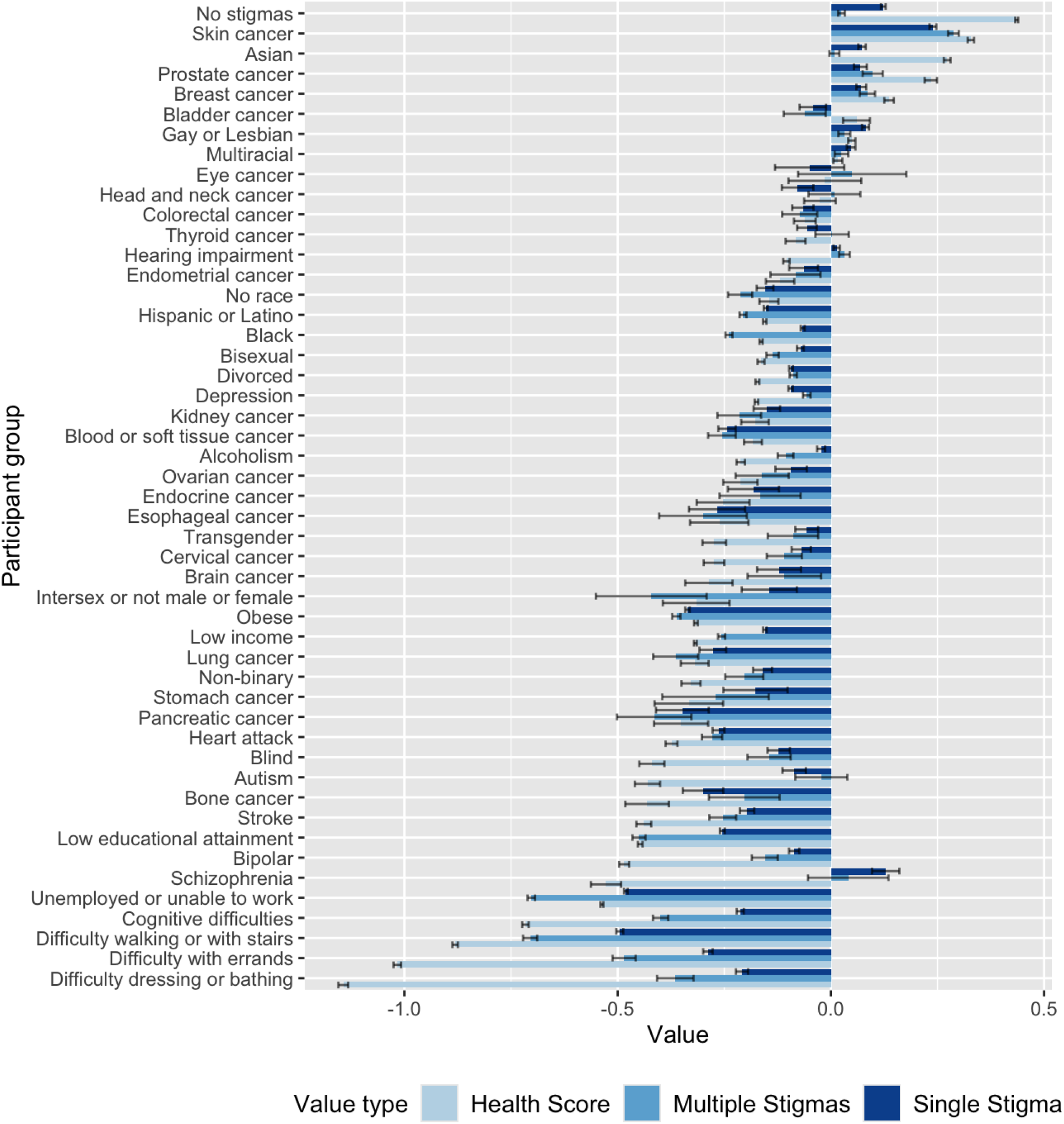
Values for the average overall health rating per single stigma relative to the mean for the cohort as a whole, and their estimated effect size in the model sorted by average overall health rating. The differences between the two values are primarily due to the model including all single and pairwise effect sizes.

Effect sizes of pairwise stigmatizations with individual effect sizes are in the same direction and of relatively large magnitudes tend to have an opposite direction pairwise effect, indicating the combined effect was less than the sum of the two individual effects. For example, the effect of having difficulty walking or climbing stairs or difficulty completing errands alone is a decrease in overall health rating; however the incremental effect of having both, in addition to the individual effects, is a positive effect on overall health. This is represented by an enrichment of blue in the off-diagonal elements in the lower left of Figure 2a and an enrichment of red in the upper right.

**Figure 2.**
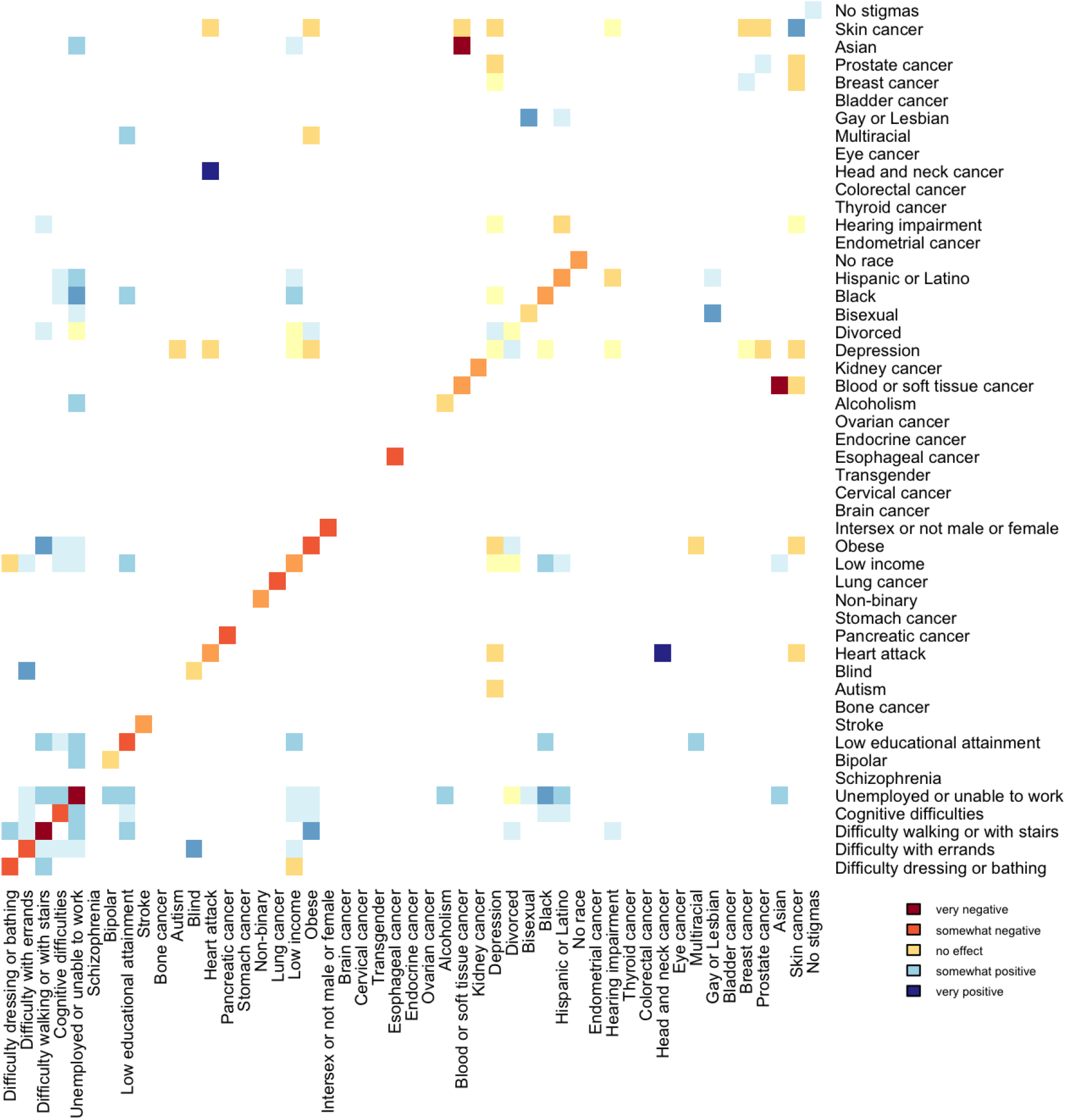
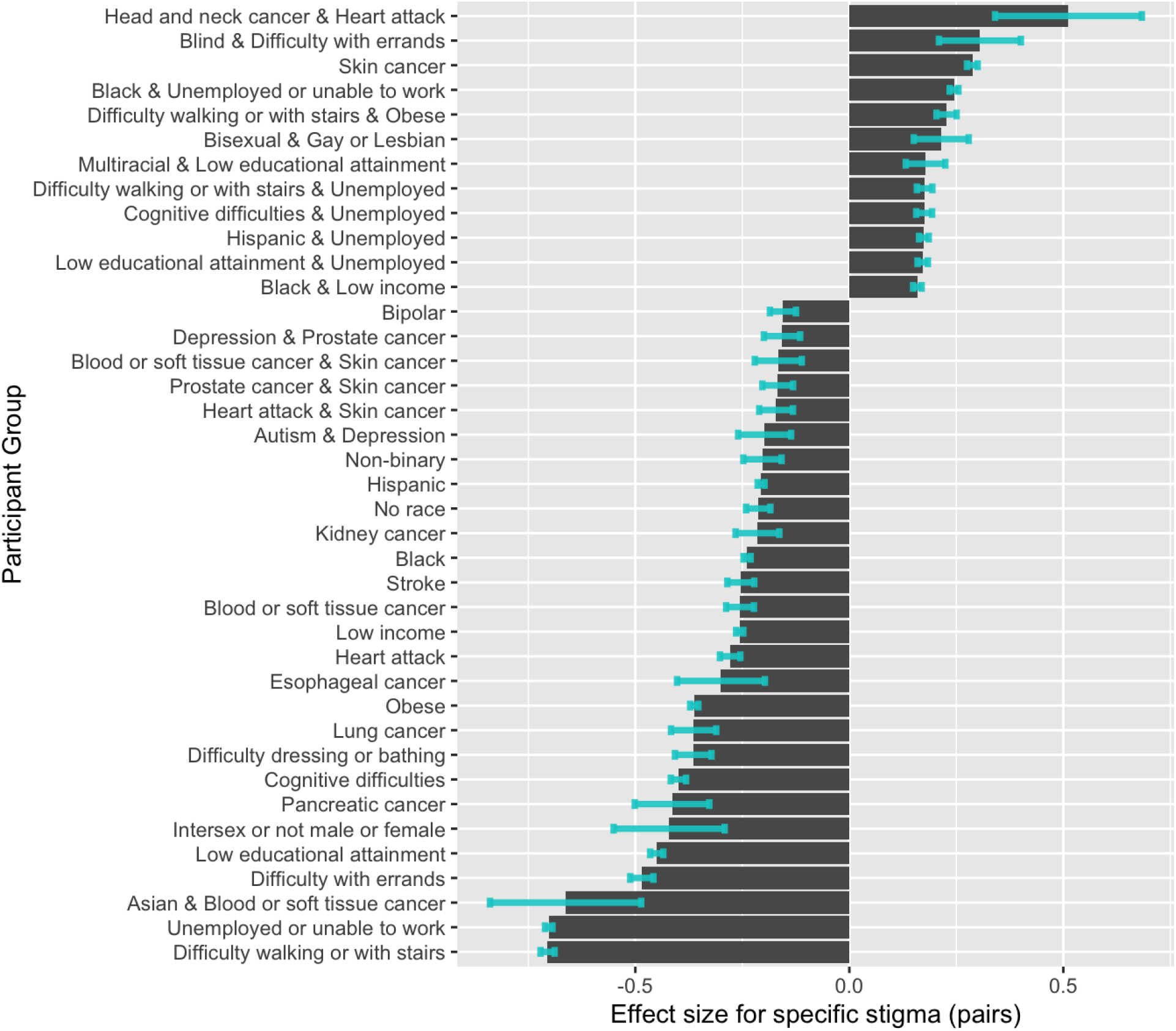
Results of the model of additive and multiplicative effects of socially stigmatized identities on self-reported overall healtha) A heat map of model estimates of the effect of individual (lower left to upper right diagonal) and pairwise (all other) stigmatized identities on self-reported overall health, sorted in the same order as in Figure 1 (i.e. based on the average overall health scores per individual identity). Red represents a negative effect on self-reported overall health, pale yellow represents neutral effect, blue represents a positive effect, and white indicates no data, due to either the pairwise stigmatizations having 20 participants or that the estimated effect is not statistically significant after using false discovery rate to correct for testing multiple hypotheses^18^. b) All individual and pairwise estimated effect sizes with absolute value of at least 0.15 and adjusted p-value lower than 0.05. Note: “Unemployed or unable to work” was shortened to “Unemployed” in this figure.

In the model that includes multiplicative effects, out of 47 individual stigmatizations, 29 had statistically significant effect sizes, as did the group that declined to report their race or ethnicity. When examining statistical significance, we used false discovery rate to adjust for testing multiple hypotheses^18^. The remaining 19 stigmatized identities had a small effect size (e.g. Asian, autism, and thyroid cancer) and/or small sample size (e.g. bladder, brain, and head and neck cancers). Except for skin cancer, all significant individual effects were negative. Out of 1,124 possible pairwise stigmas, 758 had sufficient sample sizes to estimate an effect size (with a requirement of at least 20) and 116 had statistically significant effects. The individual stigmas with the largest number of statistically significant stigma pairs were: unemployed or unable to work (14); depression and low income (11 each); and difficulty walking or climbing stairs, cognitive difficulties, obesity, and skin cancer (8 each). This is influenced by sample size; the correlation between sample size and number of statistically significant multiplicative effects is 0.78. Depression had the largest number of negative pairwise effect sizes, indicating it negatively modifies other stigmatizations, alongside its minor negative effect alone.

The pair with the largest negative estimated effect (Figure 2b) is Asian and blood or soft tissue cancer, which is consistent with prior data showing that head and neck angiosarcomas are more common in Asian populations and may have unique transcriptomic signatures ^19^. Most other pairs with negative effects are cancers, combined with other types of cancer or heart attack, along with autism and depression, and prostate cancer and depression. The pairs with the largest positive estimated effect sizes are head and neck cancer and heart attack along with blind and difficulty with errands. In pairs with a positive effect, most individual stigmas have negative effect sizes, indicating that the combined effect is less negative than the sum of the two individual effects.

## Discussion

These analyses demonstrate statistically significant effects of 29 individual stigmatized identities and 116 pairs of stigmatized identities on an individual’s rating of their overall health. They reveal a wide variety of unknown causes to be further studied, as well as pairs of intersecting variables that should be included within the analyses of specific diseases to ensure that subgroup effects are accurately characterized.

While it can be argued that each stigmatization has both direct and indirect effects on health, the relative importance of direct and indirect effects will vary. For example, participants with pancreatic cancer most likely rate their overall health as low due to the direct effects of pancreatic cancer, whereas individuals with low income likely rate their overall health lower due to indirect effects such as reduced access to healthcare ^20^. The large negative estimated effect of low educational attainment is striking and indicates a need to redouble efforts to improve health literacy within this population.

Cultural components of self-rated overall health should be explored further to understand how they affect health care. For example, prior work has noted higher overall health self-ratings from Asian participants compared to white participants with similar characteristics ^21^, which correlates with the higher overall health scores observed here (Figure 1).

This work has limitations that apply to many efforts to quantify intersectionality ^22^ including 1) being limited to studying the stigmatizations for which there was sufficient data available; half of the stigmatized statuses were not able to be included ^17^; 2) that whether each participant has a specific status is based on one point in time, many statuses change over time, such as mental health diagnoses, disability statuses, and employment status; 3) the encoding of all stigmatizations as binary variables, even those for which a larger domain could be used (e.g. educational attainment and income); 4) that the data were collected from participants only in the United States; and that only individual and pairwise stigmatization was considered, when there are likely higher order effects that could not be analyzed while maintaining adequate statistical power. These limitations can be addressed in future research, for example by studying smaller subsets of intersectionality in greater depth. Future analyses can also delve deeper into aspects of health that vary between stigmatizations (e.g. physical vs mental health), as well as other causes of health disparities such as discrimination, access to health insurance, and access to care providers that share their race and/or ethnicity, and/or have cultural competence ^23,24^.

There are numerous pairs of stigmatized identities that significantly affect self-reported overall health and therefore should be considered in research and clinical care. Considering intersectionality when evaluating each individual and cohort’s overall health can help researchers and clinicians reduce mistakes.

## Data Availability

The data are available through the All of Us Research Program's Researcher Workbench. Code and interim analysis files are available as supplemental data.

https://researchallofus.org/

## Acknowledgements

The authors are grateful to the participants of the *All of Us* Research Program, whose generous contributions made this work possible.

## Supplemental Figures

**Figure S1.**
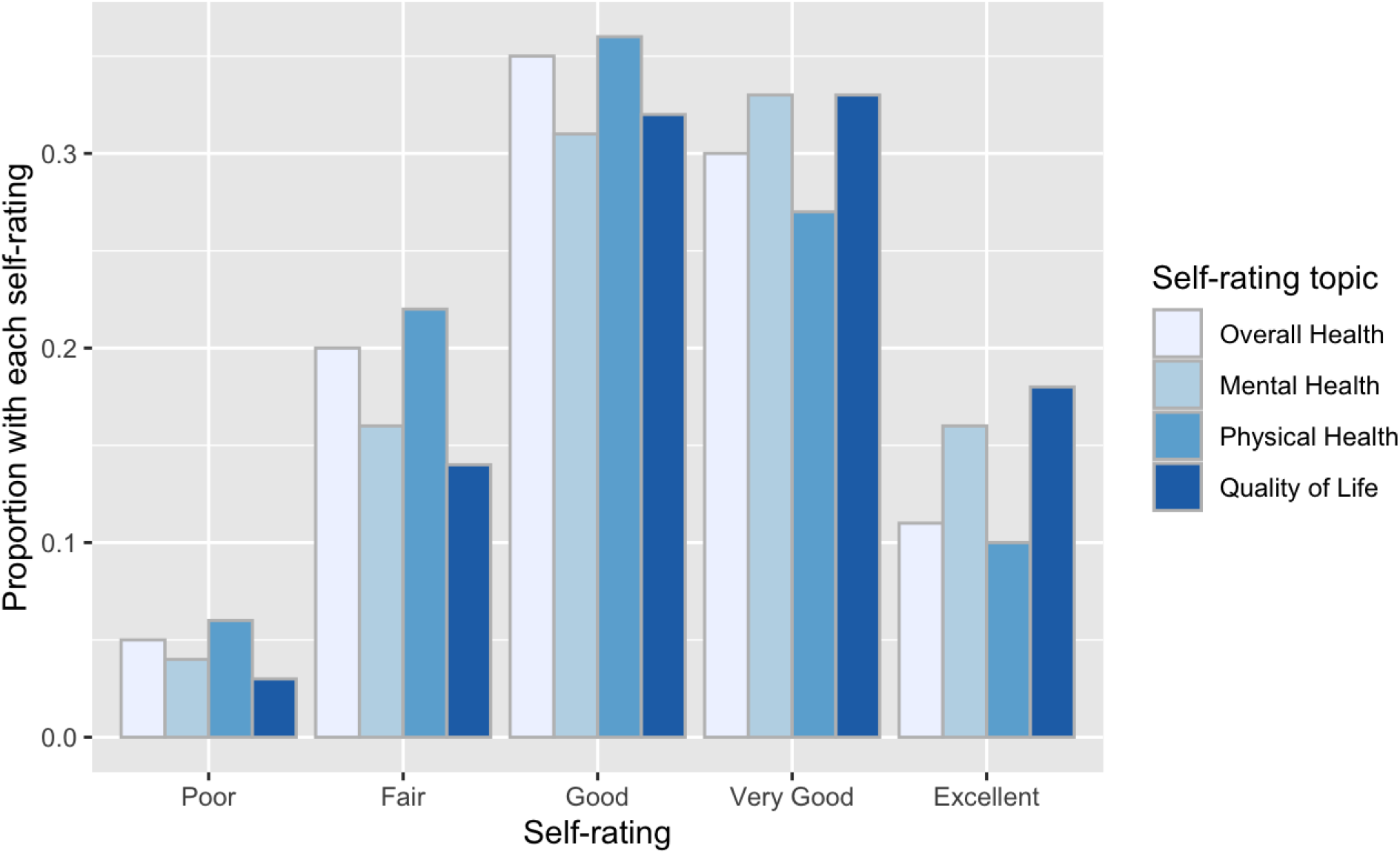
Self-ratings across dimensions of health. Because of the correlation between values (average 0.62), and the decrease in power that would have resulted from quadrupling the number of variables, we included only overall health in the model.

**Figure S2.**
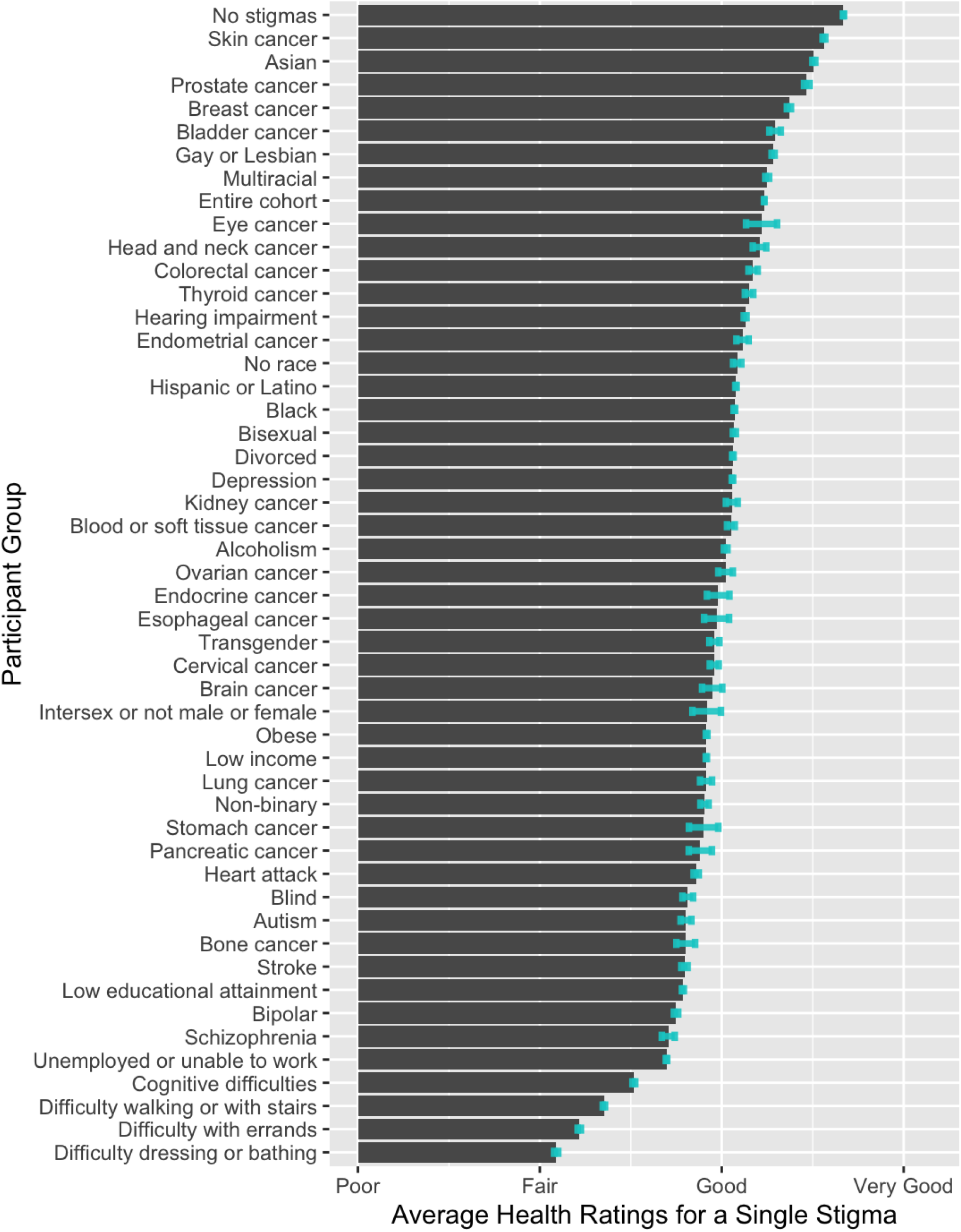
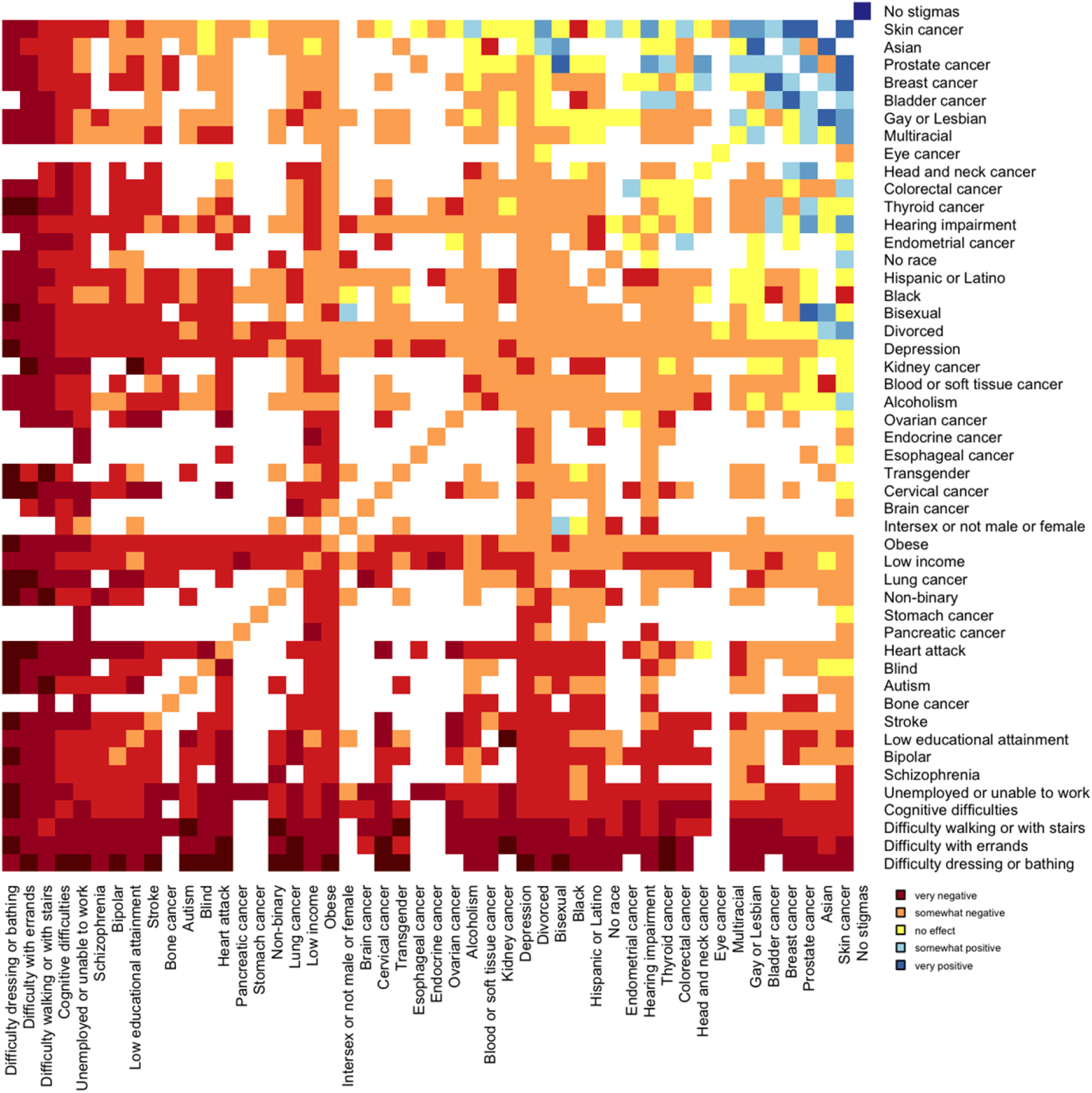
Average overall health ratings. Figure S2a: The average overall health rating and standard error of the mean of each individual participant group, sorted by estimated mean. Note, there is an option “Excellent” that is not listed because it is a larger value on the horizontal axis than any of the estimated means. Figure S2b: The average overall health rating for individual and pairwise stigmatizations, sorted by the estimated individual mean (visualized on the diagonal). Pairs with fewer than 20 participants are excluded, represented by white (as opposed to the pale yellow representing minimal effect relative to the average).

**Figure S3.**
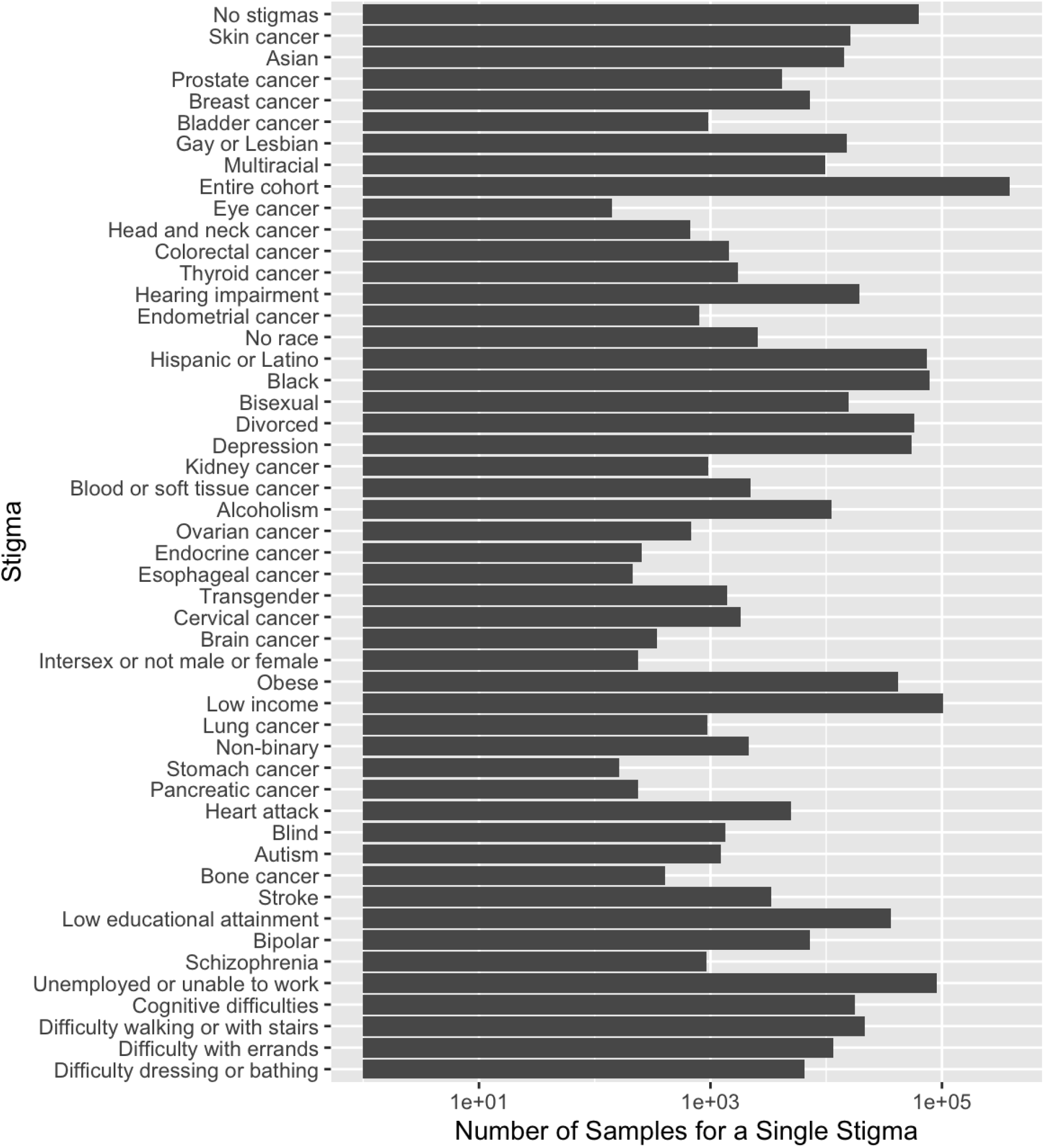
The number of participants with each individual stigma and the cohort as a whole; note the log scale. When comparing this to Figure 1, it is clear that stigmatizations with larger standard error of the mean generally have lower sample sizes.

